# G protein subunit gamma 4 expression has potential of detection, prediction, and therapeutic target for liver metastasis of gastric cancer

**DOI:** 10.1101/2020.08.14.20175034

**Authors:** Haruyoshi Tanaka, Mitsuro Kanda, Takashi Miwa, Shinichi Umeda, Koichi Sawaki, Chie Tanaka, Daisuke Kobayashi, Masamichi Hayashi, Suguru Yamada, Goro Nakayama, Masahiko Koike, Yasuhiro Kodera

## Abstract

Liver metastasis of gastric cancer is the most common for hematogenous metastases and so fatal, that the identification of novel markers and targets for therapy are crucial. We conducted transcriptome analyses between synchronous liver metastasis, primary tumor, and adjacent tissues from four patients with metastasis confined to the liver to discover that *GNG4* upregulated substantially in primary gastric cancer tissues. Quantitative RT-qPCR assay for 300 gastric cancer patients revealed that higher levels of *GNG4* in primary cancer were associated with shorter overall survival and a higher risk of liver recurrence. The oncogenic phenotypes of *GNG4* were determined by knockout and forced expression of *GNG4*. Tumor formation by *GNG4* knockout cells was more strikingly attenuated in a liver metastasis mouse model compared with a subcutaneous model. *GNG4* is a candidate for a therapeutic target for liver metastasis, and its expression may enable us to provide better disease monitoring for liver metastasis.

## INTRODUCTION

Gastric cancer represents the third most leading cause of death among malignancies (Allemani et al., 2018; Ferlay et al., 2019; Tan & Yeoh, 2015). The 5-year overall survival, varying from 20% to 50%, remains dismal, and its trends improved little or rather flat in this decade (Allemani et al., 2018). At early stages, most instances of gastric cancer can be eradicated surgically (Hiki et al., 2018). What worsens outcomes in advanced gastric cancer is distant synchronous and metachronous metastases. To minimize the risk of metachronous metastases, gastrectomy with perioperative chemotherapy or chemoradiotherapy are standard therapies. Although several molecular targeted drugs also have become available, they are only recently being considered for their indications (Bang et al., 2010; Kang et al., 2017; Shitara et al., 2018).

The liver is the most common site of hematogenous metastasis from gastric cancer, yet the diagnostic and therapeutic strategy for treating liver metastasis remains demanding (Kodera et al., 2014). Although palliative therapy is standard care for liver metastasis, it is getting considered that single lobe hepatectomy with R0 margin for a solitary nodule can improve their prognoses for either synchronous or metachronous liver metastases (Kodera et al., 2014; Markar et al., 2016; Van Cutsem et al., 2016). Despite this trend, chemotherapeutic strategies for liver metastasis are still limited to the same general protocol as those used for other distant metastasis, unlike specific therapies available for peritoneal metastasis (Ishigami et al., 2018; Takahashi et al., 2018). It is also challenging to diagnose small metastasis in liver accurately preoperatively. Moreover, early disease recurrence may imply subclinical dissemination has occurred at the time of surgery (Amikura et al., 1995; Sakamoto et al., 2020).

Cancer cells of the primary gastric tumor are considered to enter circulation through the portal vein to the liver to form metastatic foci. The background of this inference comes from the ‘seed and soil’ theory. This theory advocates during the process of every metastasis, cancer cells acquire diverse malignant phenotypes through various selective pressure according to organs to which cancer cells to metastasize (de Groot et al., 2017; Mathot & Stenninger, 2012; Mendoza & Khanna, 2009). Once these cancer cells detach from the primary foci and enter the portal vein circulation, they are exposed to hypoxia and anoikis (Shimizu et al., 2018). Eventually, only a subclone with suitable attributes for a microenvironment of the liver can grow there. During the last decades, bioinformatics analyses using next-generation sequencing or microarray techniques have illustrated a genetic profile of metastatic potential, and this has enabled the development of novel drugs (Brosnan & Iacobuzio-Donahue, 2012; Kim et al., 2015; Sakamoto et al., 2020; Tanaka et al., 2016). Given this achievement, there must be a novel gene profile that drives cancer cells to form metastasis specific to liver, but this gene profile is yet to be elucidated (Bang et al., 2010; Menard et al., 2003; Slamon et al., 2001).

Hence, discovering novel genes accounting for liver metastasis by organ-oriented approach will shed light on strategy for advanced gastric cancer from novel aspects. We hypothesized that primary cancer must have the same gene profile as the metastatic liver foci have, and it could provide a clue to discover a novel driver oncogene. In this study, we used a transcriptomic approach to identify the nucleotide-binding protein (G protein) subunit gamma-4 gene *(GNG4)* as a representative candidate driver of liver metastasis. In the literature, *GNG4* was only reported as a tumor suppressor gene in glioblastoma and renal cell carcinoma (Maina et al., 2005; Zhang et al., 2018). This study revealed its malignant roles and impact on gastric cancer patients’ outcomes through expression and functional studies of *GNG4*. To the best of our knowledge, this is the first study to have revealed novel malignant roles for *GNG4* in metastasis and outcomes in gastric cancer.

## MATERIALS AND METHODS

### Sample collection

For transcriptome analysis, we used 12 surgically resected specimens consisting of three foci from four patients each; primary gastric cancer tissues, normal mucosae, and liver metastases obtained from four patients of gastric cancer confined to synchronous liver metastasis (Figure 1A). To validate contribution and determine the clinical significance of *GNG4* expression, we acquired primary gastric cancer tissues with Union for International Cancer Control (UICC) stages I to IV from 300 patients who underwent gastric resection at our institution between 2001 and 2014. This cohort was independent of four patients for which we conducted transcriptomic analysis above. They did not undergo preoperative treatment. Since 2006, all patients with stage II to III gastric cancer underwent adjuvant chemotherapy using S-1 unless the patient’s condition denied indication. To add validation for the results from our institutional data, we employed the GSE62254 dataset, global gene expression experiments (Cristescu et al., 2015; Szasz et al., 2016). We acquired a cell line MKN1 from the American Type Culture Collection (Manassas, VA, USA) for *in vitro* and *in vivo* experiments. All informed consent for the use of clinical samples and data was obtained from all patients in written manners, consistent with the requirement Institutional Review Board at Nagoya University, Japan.

**Figure 1.**
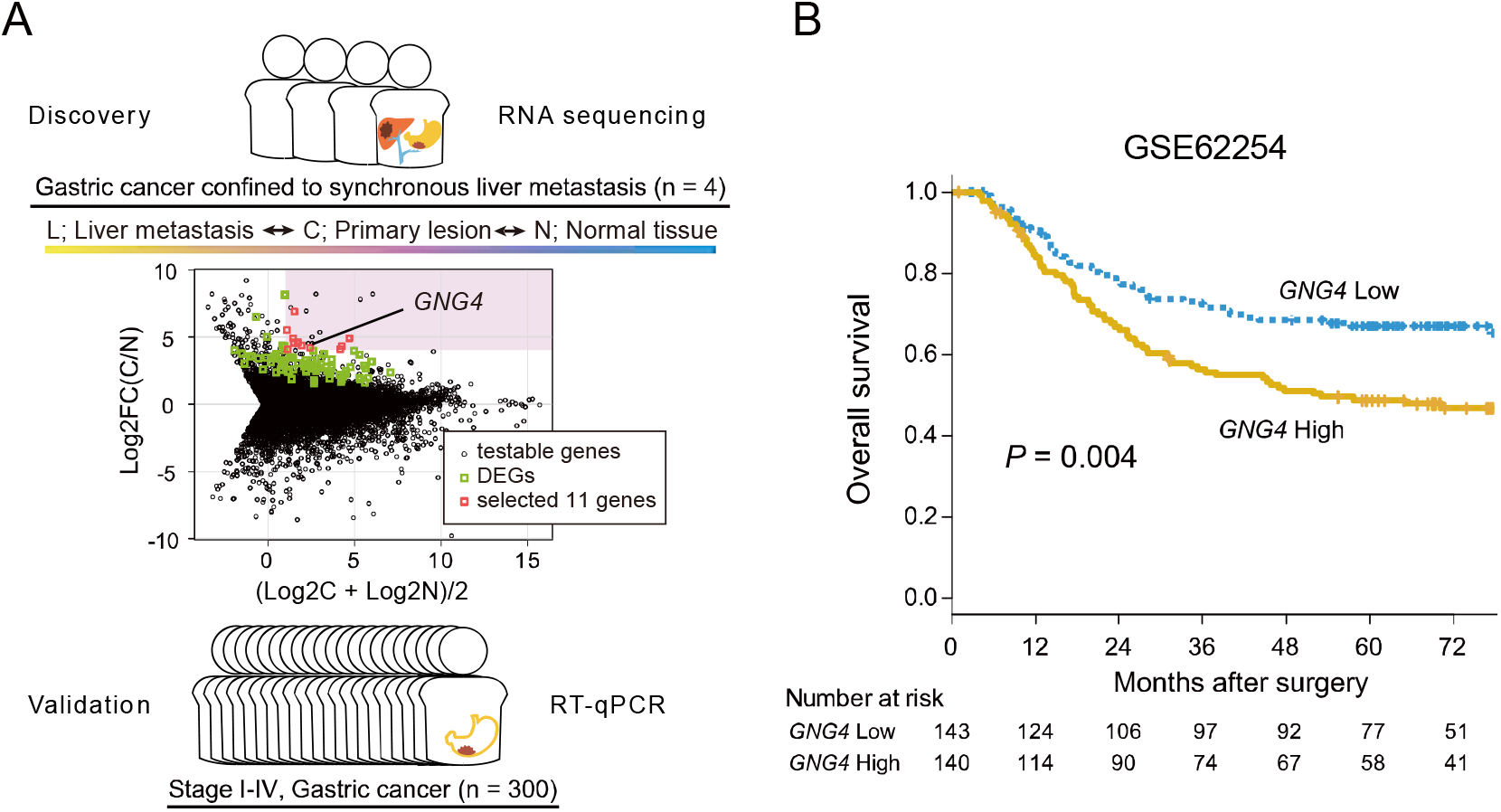
*GNG4* is identified as a putative oncogene driving liver metastasis of gastric cancer. Present study design is illustrated. 57,749 genes from RNA-seq analysis on 12 specimens from gastric cancer patients with metastasis confined to the liver. Log-ratio and mean average (MA) plot of expression levels of C and N to select putative eleven genes (A). DEGs, differentially expressed genes. Kaplan-Meier plot of overall survival using GSE62254 dataset stratified by the median value of*GNG4* (B).

### Gene expressional analyses

For global gene expression profiling analysis, we applied HiSeq Sequencing System (Illumina, San Diego, CA) to compare the expression levels of 57,749 genes as described previously (Tanaka et al., 2018). We used quantitative RT-PCR (qRT-PCR) to determine *GNG4* mRNA levels using previously published protocols (Kanda, Tanaka, et al., 2016). The primer sequence for qRT-PCR specifically designed for *GNG4* is provided in Supplemental Table 1. Immunoblot assays were performed using a rabbit anti-*GNG4* monoclonal antibody (Product code, 13780-1-AP; Proteintech, Rosemont, IL, USA), as previously described (Oya et al., 2016). Wess (Protein Simple, San Jose, CA, USA) was used for pathway analyses by immunoblotting.

**Supplementary Table 1.**
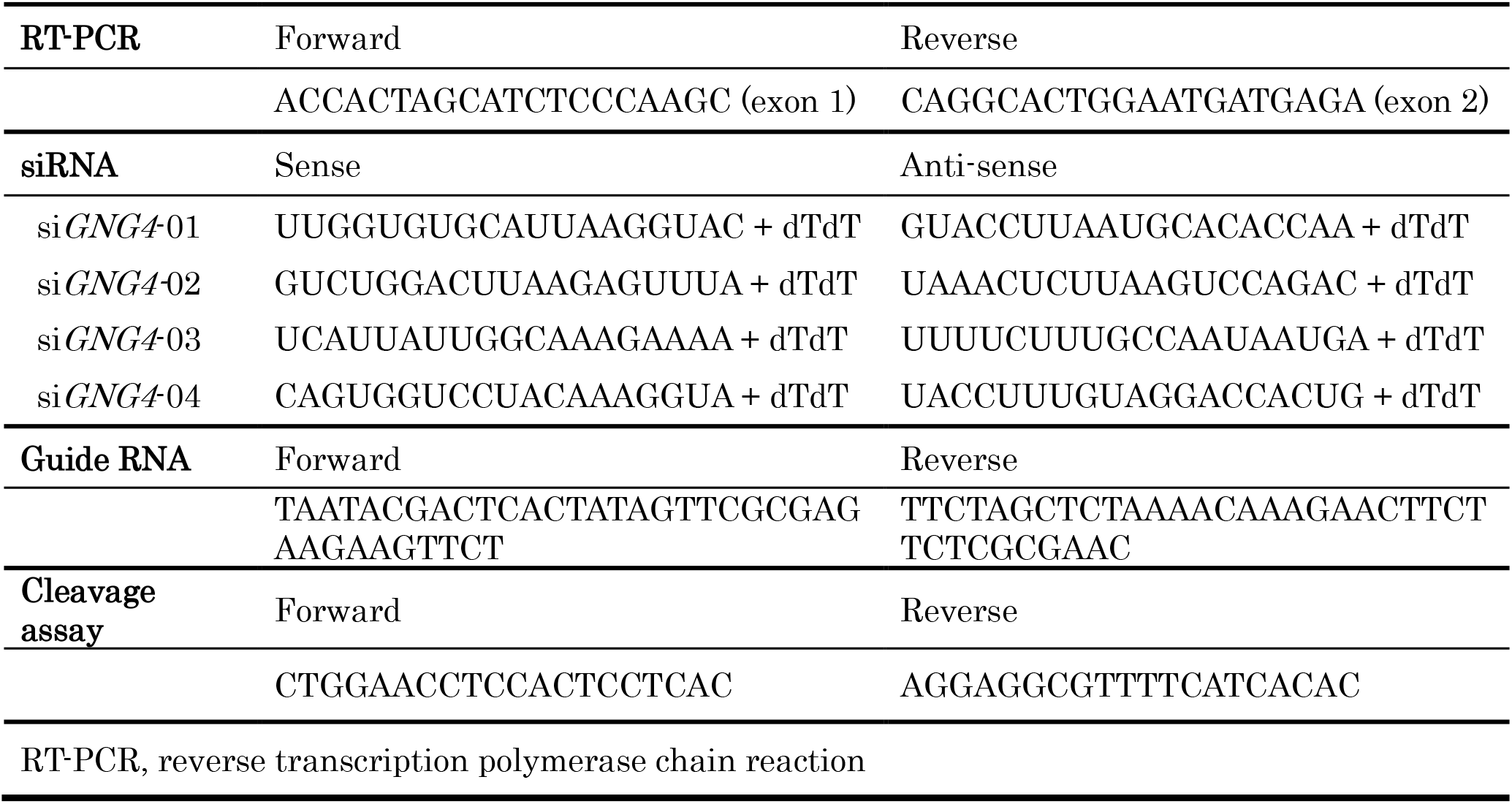
Primer sequences

### Knockout and forced expression of GNG4

We performed genome editing applying the CRISPR/Cas9 system to generate stable GNG4-knockout (dGNG4) gastric cancer cell lines, as previously reported (Kanda et al., 2018). By the GeneArt Genomic Cleavage Detection Kit (Thermo Fisher Scientific, Waltham, MA, USA), the efficiency of genomic cleavage was evaluated 72 hours after the transfection of Cas9. The primer set is described in Supplemental Table 1.

For forced expression of *GNG4*, the *GNG4* cDNA clones ligated as open reading frame sequences into CMV Flexi Vector pFN21A (GenBank ID: EU621374.1) were purchased (product ID, FHC02107; Promega, Madison, WI, USA). Using the NEON® (Thermo Fisher) system, 0.2μg of the *GNG4* vector was transfected into MKN1cells (1 x 10^5^). Analyses of phenotypes were performed from the second day of transfection,

### Functional assays and mouse models of gastric cancer

Cell proliferation was evaluated using the Premix WST-8 Cell Proliferation Assay Kit (DOJINDO Inc., Kumamoto, Japan), as previously described (Shibata et al., 2017).Distributions of cells in specific phases of the cell cycle were evaluated applying the Cell Clock Cell Cycle Assay (Biocolor Ltd, Antrim, United Kingdom). To detect apoptotic cells with depolarized mitochondrial membranes, Mito Potential Kit (Merk, Darmstadt, Germany) was used through Muse Cell Analyzer (Merck) (Kanda et al., 2018). We tested drug sensitivity analysis for 5FU, using WST-8 kit, as described previously (Kanda, Shimizu, et al., 2016). Adhesion of gastric cancer cells to a solid matrix was evaluated by the CytoSelect 48 Well Cell Adhesion Assay Kit (Cell Biolabs, San Diego, CA, USA) as previously described (Miwa et al., 2017).

To evaluate the influence of *GNG4* on tumorigenicity *in vivo*, at the outset, we employed a mouse subcutaneous xenograft model as previously reported (BALB/c-nu/nu; n = 3, each) (Kanda et al., 2018). We evaluated the role of *GNG4* in liver metastasis using an already established mouse model (NOD-SCID; n = 4, each) (CLEA Japan Inc., Tokyo, Japan) (Kanda et al., 2018; Miwa et al., 2019). The IVIS Spectrum imaging system (Xenogen, Alameda, CA, USA), and magnetic resonance imaging (MRI) (MRS 3000, MR solutions, Guildford, UK) were employed to observe tumor formation in the livers of engrafted mice. All animal experiments were conducted consistent with the ARRIVE guidelines and were approved by the Animal Research Committee of Nagoya University.

### Statistical analysis

The significance of differences between the two variables was assessed using the Student t-test for normally distributed data and the Wilcoxon test for non-normally distributed data. Fisher’s exact was used to analyze the significances of categorical data. *P* values < 0.05 were considered statistically significant. For multiple comparisons, *P* values were adjusted by the Bonferroni-Hochberg method. For time-survival analyses, survival curves were generated using the Kaplan-Meier method, and Cox proportional hazards models were used to estimate the hazard ratio and *P* values. Analyses method of transcriptomic analysis were described previously (Tanaka et al., 2018). All other statistical analyses were performed using R version 3.6.1 software (The R Foundation for Statistical Computing, Vienna, Austria).

## RESULTS

### Identification of GNG4 as a candidate driver gene of liver metastasis of gastric cancer

Global expression profiling was conducted to compare 5 the expression levels of 57,749 genes between primary lesion, liver metastasis, and adjacent normal tissues. The qualities of the RNA preparations were sufficient for analysis as indicated follows: yield data per sample = 2,981 Mb, mean reads per sample = 29,512,162 pairs, mean rate ≥ Q30 = 94.40%, mean quality score = 34.05, and mean total mapped-read rate = 89.38%. The 57,749 genes were filtered to 10 yield 94 genes according to following five conditions: (1) a significant difference (*Q* < 0.05) between primary cancer lesions (C) and adjacent normal gastric tissues (N), (2) upregulated in C, and (3) not significantly different (*P* ≥ 0.05) between C and liver metastatic lesions (H). We next selected 11 genes by filtering the 94 overlapping genes according to (4) the base-2 logarithm of fold change between C and N expression levels [log2FC(C/N)] > 4 and (5) the 15 average expression levels of C and N [(log2C + log2N)/2] >1 (Figure 1A, Table 1)

**Table 1.** Differentially expressed genes associated with liver metastasis.

| Gene symbol | Chromosomal locus | Function | C/N |  | L/C |  |
| --- | --- | --- | --- | --- | --- | --- |
|  |  |  | Log2FC | *Q* | Log2FC | *P* |
| *GNG4* | 1q42.3 | Signal transducer | 4.84 | 0.005 | 0.29 | 0.73 |
| *TNFRSF11B* | 8q24.12 | Apoptosis modulator | 4.57 | 0.005 | 0.53 | 0.43 |
| *VGF* | 7q22.1 | Growth factor | 5.46 | 0.005 | -0.81 | 0.34 |
| *DNAJC12* | 10q21.3 | Protein modulator | 4.15 | 0.005 | -1.16 | 0.10 |
| *RBP4* | 10q23.33 | Retinol carrier | 4.25 | 0.005 | 1.51 | 0.05 |
| *SYT7* | 11q12.2 | Signal transducer | 4.29 | 0.005 | 0.30 | 0.63 |
| *NPY* | 7p15.3 | Signal transducer | 4.86 | 0.005 | 0.09 | 0.90 |
| *THBS4* | 5q14.1 | Adhesion modulator | 4.01 | 0.005 | 0.95 | 0.28 |
| *KRT6B* | 12q13.13 | Structural molecule | 6.83 | 0.034 | -1.53 | 0.08 |
| *FNDC1* | 6q25.3 | Signal transducer | 4.50 | 0.005 | -0.89 | 0.16 |
| *UTS2R* | 17q25.3 | Signal receptor | 4.50 | 0.005 | 0.50 | 0.57 |

C, primary cancer tissue; N, adjacent normal tissue; L, liver metastasis; FC, fold change

We chose *GNG4* for further analysis as a candidate driver gene that promotes liver metastasis, as GSE62254 dataset revealed that higher levels of *GNG4* expression in gastric cancer tissues were associated with shorter overall survival (hazard ratio 1.73 [95% confidence interval, 1.20-2.50], *P* = 0.004) (Figure 1B). Being able to reach little information on *GNG4* (Pal et al., 2016), we identified this gene as a potential candidate for a therapeutic target and a biomarker for liver metastasis of gastric cancer and brought it further analysis.

### Clinical implications of GNG4 expression in tumor tissues

First, looking at the *GNG4* mRNA expression levels in cancer tissues among stages I-IV disease, we found that *GNG4* expressions of stage IV were significantly higher than stage I and II-III, and there was no significant difference between stages I to III (Figure 2A). Among stage IV, patients with liver metastasis had higher *GNG4* levels than without it. However, this was not the case for peritoneal and nodal distal metastasis (Figure 2B).

**Figure 2.**
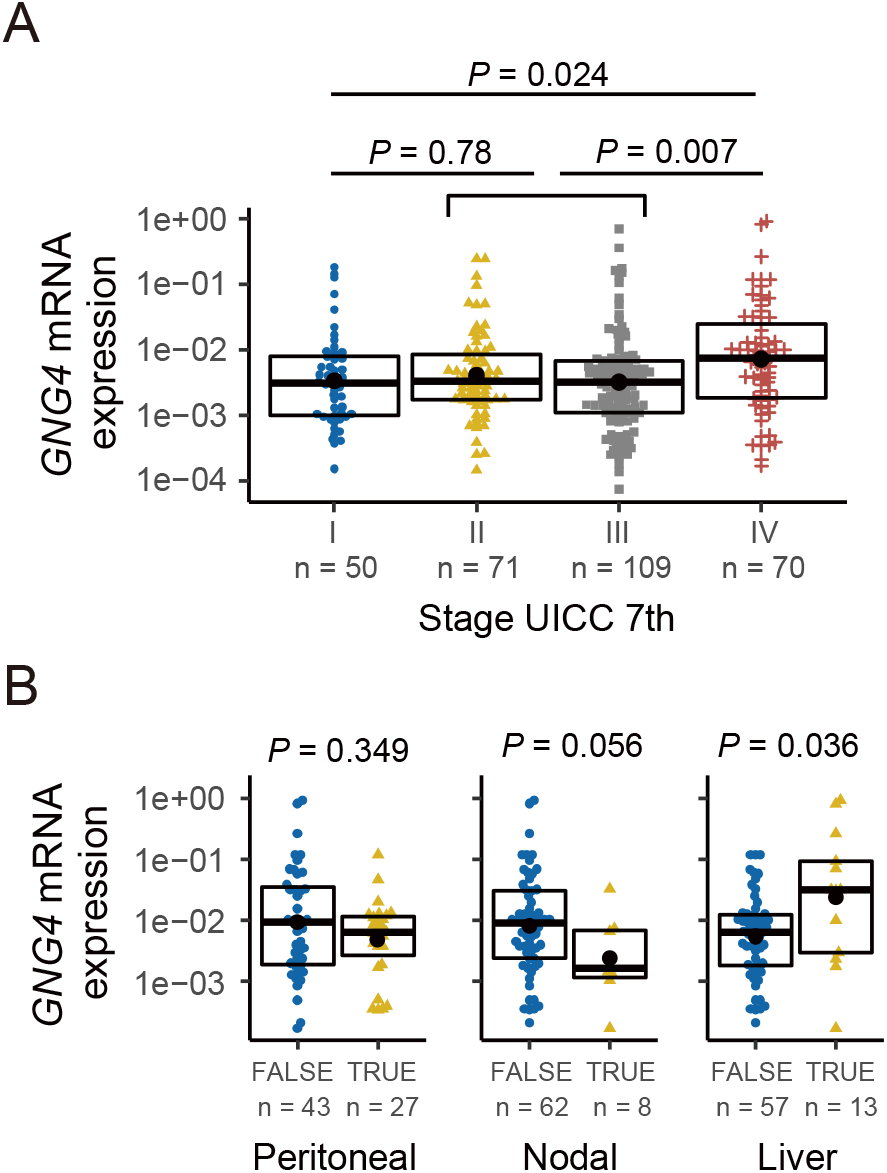
GNG4 expression was high in among UICC stage IV gastric cancer tissues, especially in with liver metastasis. GNG4 mRNA expression (GNG4/GAPDH) stratified by the UICC stage (stage I vs. II+III vs. IV) (A) and with or without (true/false) three distant metastatic foci (peritoneal, lymph node, liver) among UICC stage IV cohort (n = 71). Box, bold bar, and black point indicates interquartile range, median, and mean, respectively.

We moved on to set the cutoff value to see the implications of *GNG4* expressions. The receiver operating characteristic 5 (ROC) curve was generated to predict liver recurrence within one year postoperatively; the area under the curve was 0.696. By using this ROC, we defined high *GNG4* expression as *GNG4*/*GAPDH* > 0.004097, according to the Youden index (Supplemental Figure 1). Consistent with the association between UICC Stages and *GNG4* expressions among the whole cohort, the high *GNG4* group had shorter overall survival significantly than the low group had (hazard ratio, 1.94 [1.32–2.85]; *P* < 0.001) (Figure 3A).

**Supplemental Figure 1.**
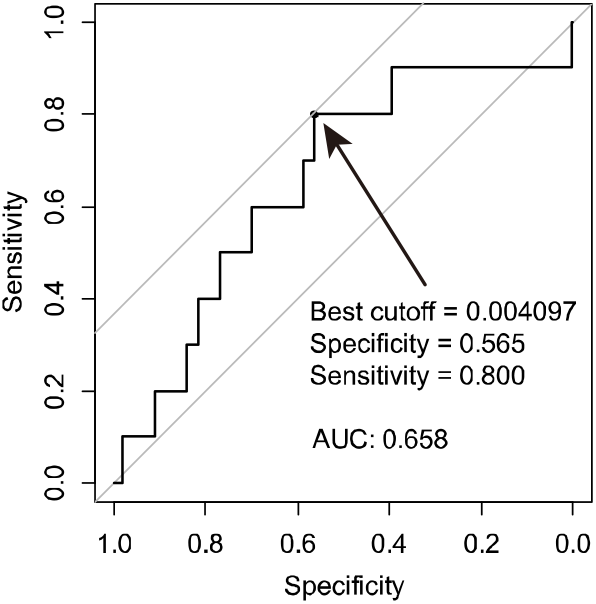
receiver operating characteristic (ROC) curve to predict liver recurrence within one year among U|CC stage II-III cohort.

**Figure 3.**
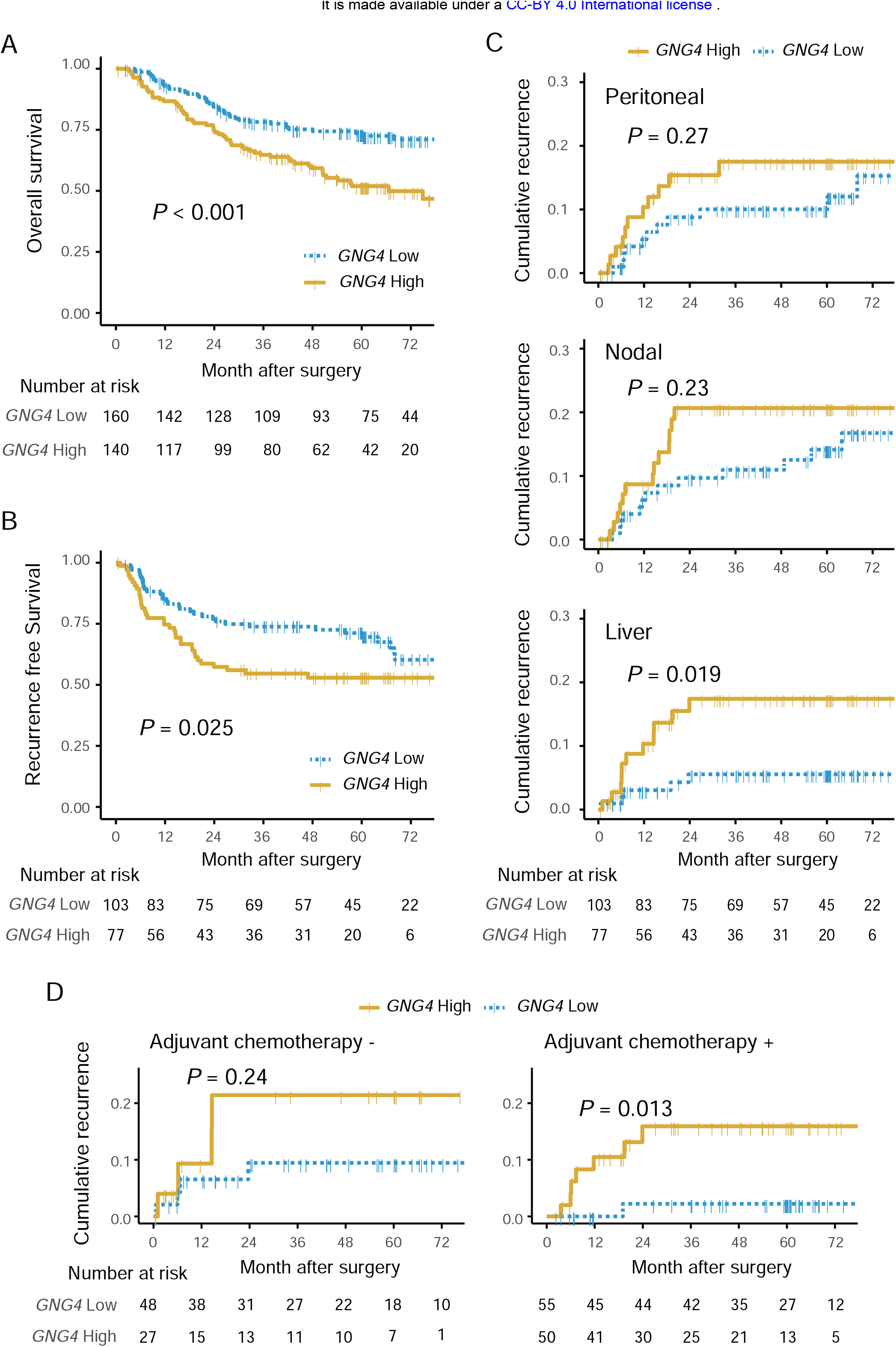
High GNG4 expression correlated with the poor survival of patients with gastric cancer. Kaplan-Meier plot of overall survival for the whole cohort (N = 300) stratified by GNG4 expression (A) and recurrence-free survival for stage II and III cohort (n = 180). Cumulative incidence stratified according to the first site of recurrence (C). Kaplan-Meier plot assessing the implications of GNG4 expression on cumulative recurrence as liver metastasis, stratified by with or without administration of adjuvant therapy (D).

We next focused on stage II-III cohort (n = 180) to see the *GNG4’s* impact on prognosis, particularly recurrence and its pattern. All of *GNG4* clinicopathological valuable similarly distributed between high and low *GNG4* expression groups, including stage II or III (Supplemental Table 2). The high *GNG4* group, nevertheless, exhibited poorer recurrence-free survival (hazard ratio, 1.73 [1.07-2.79]; *P* = 0.025) (Figure 3B). The cumulative recurrence rate for liver metastasis was significantly higher in the high *GNG4* group compared to the low group (hazard ratio, 3.34 [1.16-9.63]; *P* = 0.019), but not a case for either peritoneal or nodal recurrence (Figure 3C). High *GNG4* expression was an only risk factor for liver recurrence independently according to multivariate analysis (hazard ratio, 3.471 [1.20-10.0]; *P* = 0.022) (Table 2). We further tested if *GNG4* represent a biomarker for chemotherapy resistance, by stratified into subsets with or without chemotherapy. Among the only subset with adjuvant chemotherapy, the high *GNG4* group showed higher liver recurrence rate significantly (hazard ratio, 8.32 [1.02-67.7], *P* = 0.013), but not significant among the subset without adjuvant chemotherapy (Figure 3D).

**Supplemental Table 2.**
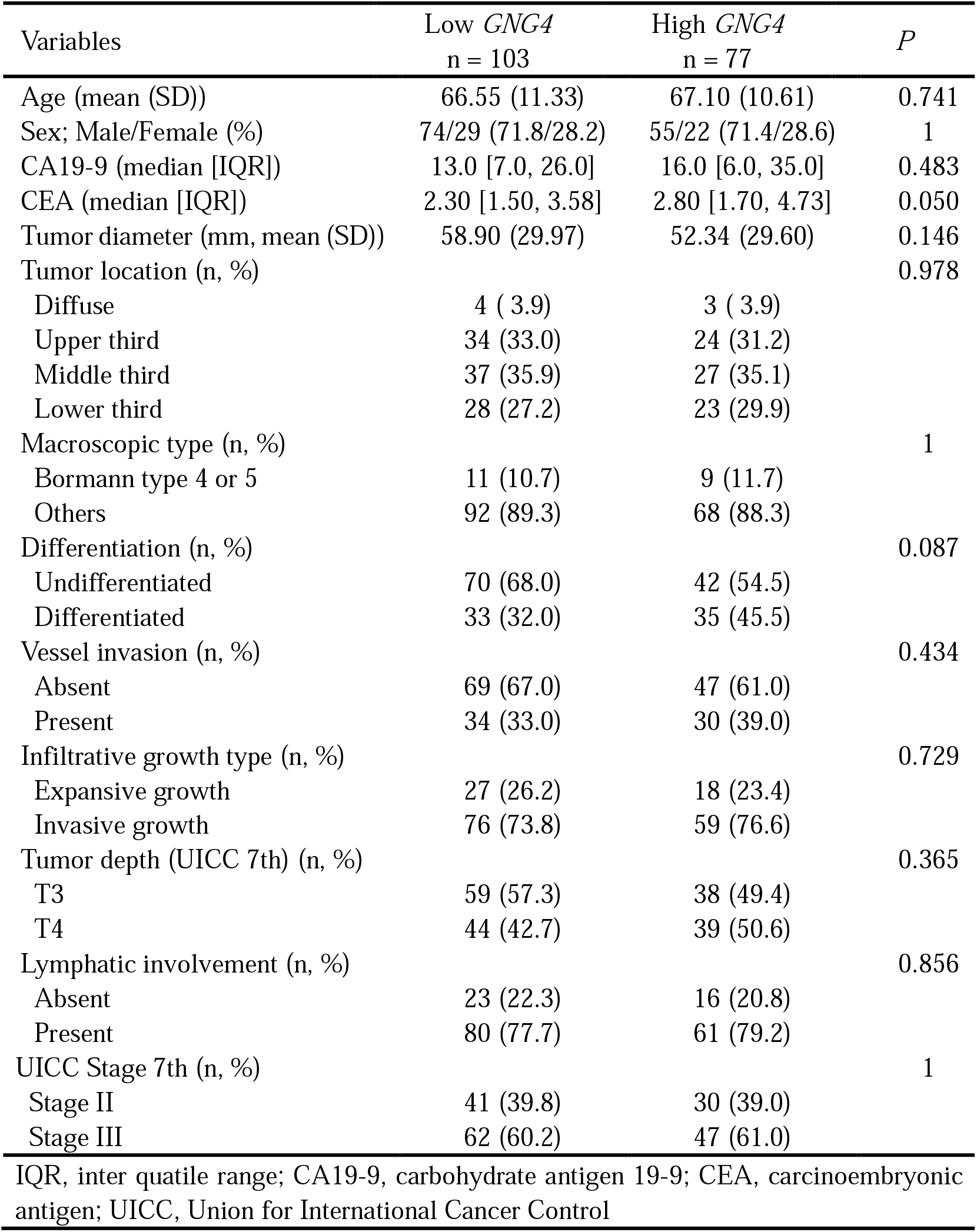
Association between high/low *GNG4* mRNA levels and clinicopathological factors of 180 patients of stage II/III patients.

**Table 2.** Predictive factors of liver recurrence for 180 patients with R0 resection of gastric cancer

|  |  |  |  |  |  |  |  |  |
| --- | --- | --- | --- | --- | --- | --- | --- | --- |
| Variables | Univariate |  |  |  | Multivariate |  |  |  |
|  | HR | 95%CI<br>Lower | 95%CI<br>Upper | *P* | HR | 95%CI<br>Lower | 95%CI<br>Upper | *P* |
| Age >65 years old | 1.59 | 0.551 | 4.57 | 0.380 |  |  |  |  |
| Male sex | 2.84 | 0.645 | 12.5 | 0.118 |  |  |  |  |
| Tumor size >50 mm | 0.96 | 0.361 | 2.57 | 0.939 |  |  |  |  |
| Tumor Location (lower third) | 1.62 | 0.605 | 4.36 | 0.343 |  |  |  |  |
| CA19-9 >37 IU/mL | 2.96 | 1.07 | 8.15 | 0.049* | 2.27 | 0.813 | 6.34 | 0.118 |
| CEA >5 ng/mL | 1.84 | 0.593 | 5.71 | 0.317 |  |  |  |  |
| Differentiated | 2.59 | 0.943 | 7.14 | 0.059 |  |  |  |  |
| Invasive growth type | 0.12 | 0.016 | 0.93 | 0.006* | 0.141 | 0.018 | 1.08 | 0.059 |
| Vessel involvement | 5.58 | 0.737 | 42.3 | 0.032* | 3.77 | 0.486 | 29.3 | 0.204 |
| Lymph node metastasis | 0.98 | 0.366 | 2.64 | 0.972 |  |  |  |  |
| Tumor depth T4 (vs T1-3) | 2.35 | 0.534 | 10.4 | 0.210 |  |  |  |  |
| UICC Stage III | 1.00 | 0.370 | 2.68 | 0.993 |  |  |  |  |
| High *GNG4* | 3.34 | 1.16 | 9.628 | 0.019 | 3.471 | 1.201 | 10.027 | 0.022* |

\*Statistically significant ( $P < 0.05$ ); CI, confidence interval; CEA, carcinoembryonic antigen; CA19-9, carbohydrate antigen 19-9; UICC, Union for International Cancer Control

### The oncogenic phenotype of GNG4 in gastric cancer cells

To see *GNG4* functional aspects, we modulated *GNG4* expression. We established a stable *GNG4* knockout *(dGNG4)* MKN1 cell line applying the CRISPR/Cas9 system. The knockout of *GNG4* was confirmed using an amplicon-cleavage assay and immunoblotting (Supplemental Figure 2A and 2B, respectively). Decreased cell proliferation was observed by knockout of *GNG4* (Figure 4A), and forced expression of *GNG4* increased MKN1 cell proliferation (Figure 4B). The phase of dGNG4-MKN1’s cell cycle appeared to have arrested in S/G2 (Figure 4C and Supplemental Figure 2C). In search of the reason for the cell-cycle arrest, we tested whether *GNG4* plays a role in maintaining mitochondrial membrane polarity to help cancer cells to survive. The cell proportion of which mitochondrial membrane depolarized increased in the knockout of *GNG4* (Figure 4D and Supplemental Figure 2D) and the inverse effect was observed by forced expression of *GNG4* (Figure 4E, and Supplemental Figure 2E). To test the possible role of chemotherapy-resistant, we performed a drug-sensitivity test. Knockout of *GNG4* modest increased sensitivity to 5FU, although it was still statistically significant (Figure 4F). The abilities of dGNG4-MKN1 cells adhering to collagen I, collagen IV were significantly reduced compared with that of parent MKN1 cells (Figure 4G). We explored pathways involve*GNG4* and genes interacting with *GNG4*. Extracellular signal-regulated kinase 1/2 (ERK1/2) was markedly, and Akt was moderately dephosphorylated in dGNG4-MKN 1 cells compared with parent MKN1 cells (Figure 4H).

**Figure 4.**
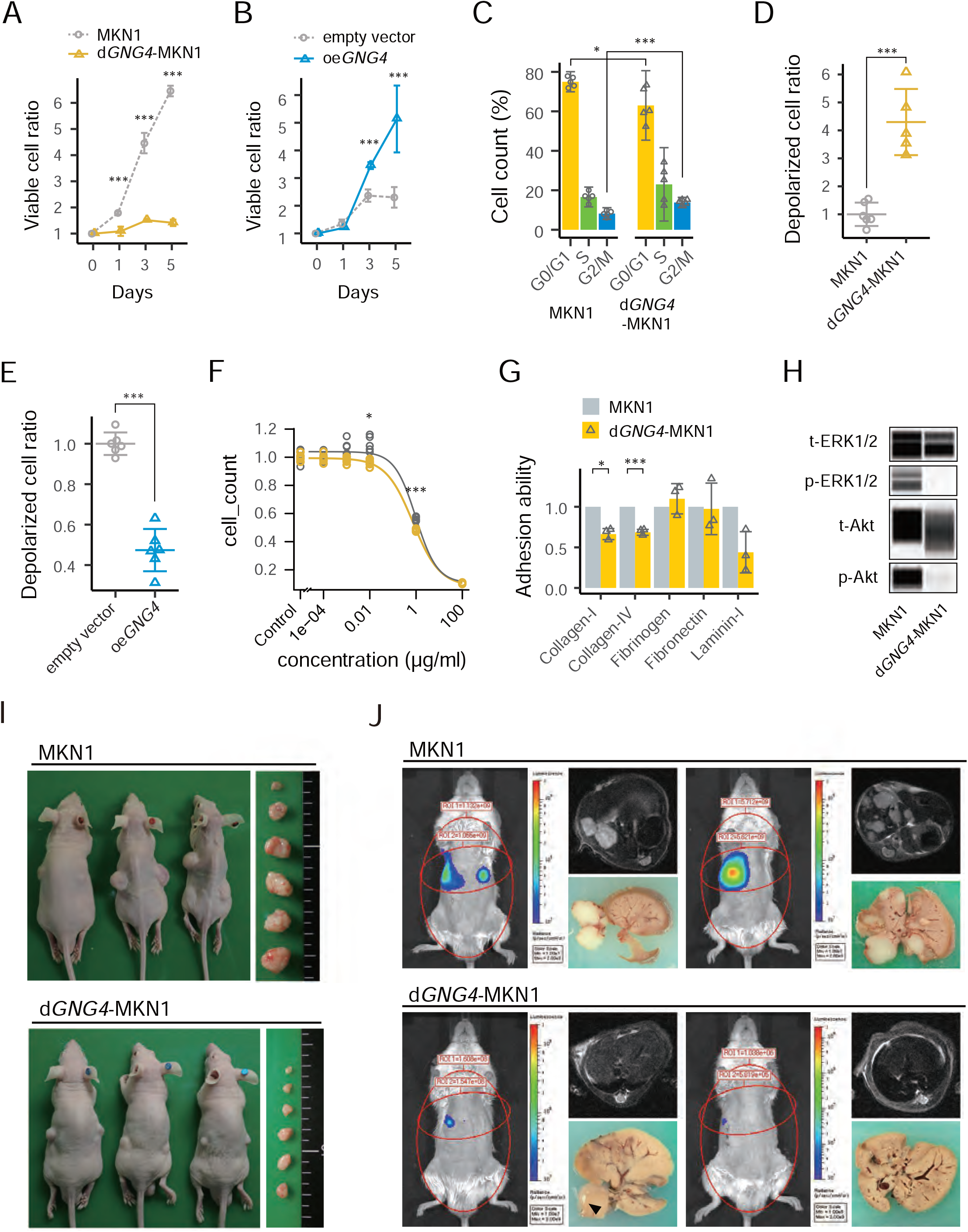
In vitro and in vivo functional assay for GNG4 determined to malignant phenotypes of MKN1 cell by knockout of GNG4 (dGNG4) and over-expression of GNG4 (oeGNG4). Proliferation assay between parental MKN1 and dGNG4-MKN1 (A), and oeGNG4-MKN1 (B). Cell cycle assay comparing MKN1 and dGNG4-MKN1 using colorimetric detection of the cell cycle phase (C). Mitochondrial membrane potential assays to identify enhanced apoptotic pathway between MKN1 and dGNG4-MKN1 (D), and oeGNG4-MKN1 (E). Drug sensitivity assay to 5-FU. The curve fitting was done by four-parameter log-logistic models (F) and cell adhesion assay to solid matrixes (G) comparing MKN1 and dGNG4-MKN1. Immunoblot assay comparing phosphorylation of Extracellular signal-regulated kinase 1/2 (ERK1/2), and Akt between MKN1 and dGNG4-MKN1(H). p-indicates phosphorylated; t-, total. Mouse subcutaneous xenograft model (I) and liver metastasis model (J) comparing MKN1 and dGNG4-MKN1. Tumor progression was evaluated by MRI and by In Vivo Spectrum Imaging System (IVIS). Circles on the mice indicate regions of interest. *P < 0.05, **P < 0.01, ***P < 0.001.

Next, we employed a subcutaneous xenograft mouse model to determine if the knockout of *GNG4*influenced the formation and tumor growth *in vivo*. The tumors generated by *dGNG4-*MKN1 cells did not grow as much as parental MKN1 cells, even stopped growing after six weeks passed from seeding. The volumes of subcutaneous tumors (= *D* x *cF/2)* generated by dGNG4-MKN1 cells were significantly smaller compared to those by parental MKN1 cells (Figure 4I, Supplemental Figure 2F). Subsequently, we employed a xenograft model of liver metastasis to determine the contribution of *GNG4* to the metastatic potential of gastric cancer cells. Metastatic nodules in the liver were multiple 12 weeks after implantation of parental MKN1 cells. In contrast, those of *dGNG4* cells were significantly smaller or not detected macroscopically, or through IVIS or MRI (Figure 4J, Supplemental Figure 2G).

**Supplemental Figure 2.**
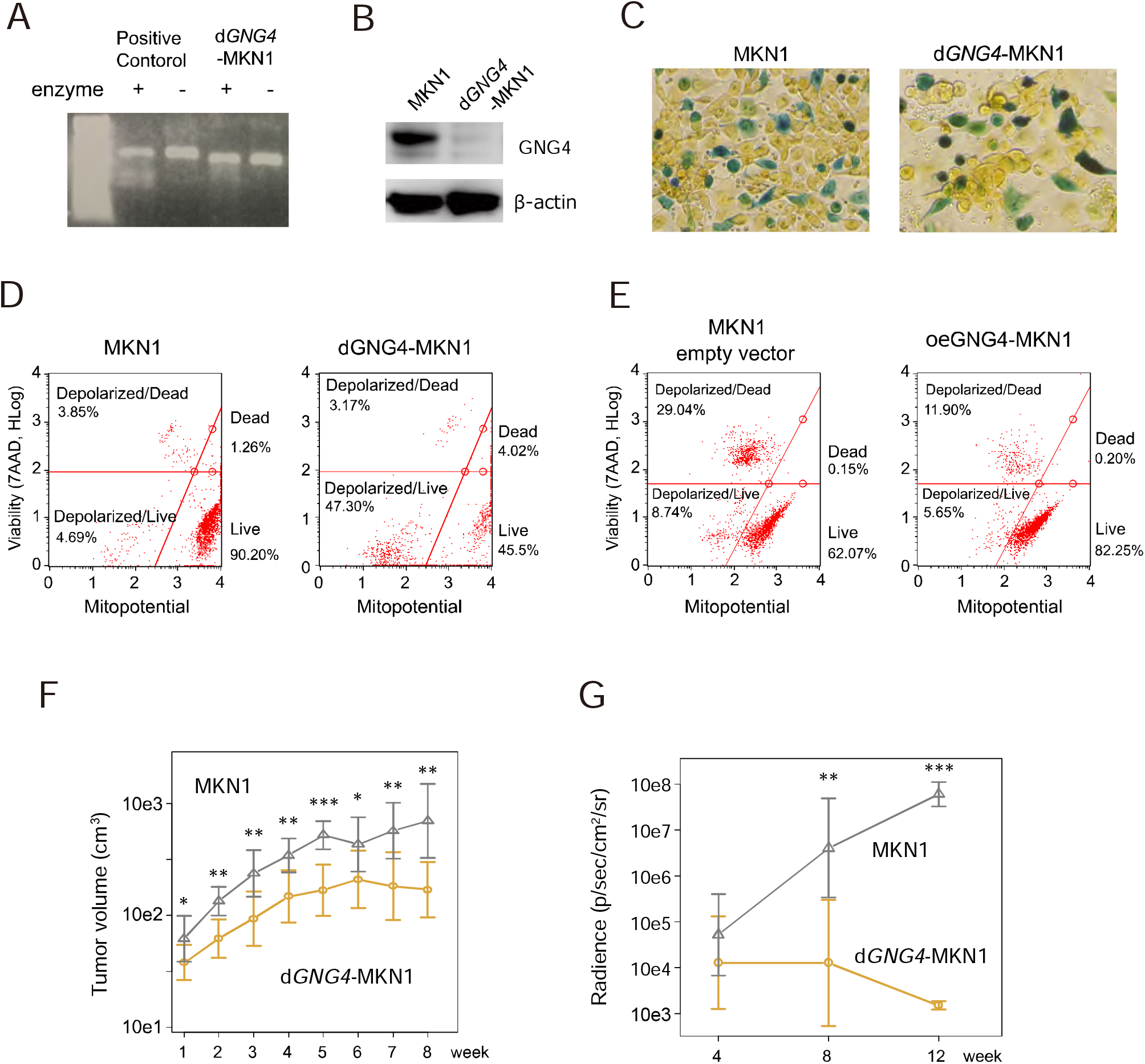
Genome editing efficacy for knockout of GNG4, using gel electrophoresis and densitometry (A), and immunoblot anlaysis of GNG4 expression (B). Colorimetric assay for detecting cell cycle phase (C). Flow cytometry detecting mitochondrial membrane depolarization (D). Tumor burden comparison between MKN1 and dGNG4-MKN1 in mouse subcutaneous xenograft model (E) and liver metastasis model (F). *P <0.05, **P < 0.01, *** P < 0.001.

## DISCUSSION

The outcome of gastric cancer remains dismal, and the diagnostic and therapeutic strategy for liver metastasis has not developed substantially (Allemani et al., 2018; Ferlay et al., 2019; Tan & Yeoh, 2015). The fact that the liver is the most common site for hematogenous metastasis inspired us to hypothesize that the same gene would overexpress at primary cancer as the metastatic liver foci. Here we identify *GNG4* as a candidate driver gene that promotes liver metastasis in patients with gastric cancer. Transcriptome analyses uncovered that *GNG4* was distinctly overexpressed in primary tumor tissues of patients with confined to synchronous liver metastases. Subsequent qRT-PCR analysis of 300 gastric cancer tissues exhibited that higher *GNG4* expression correlates with a worse prognosis, particularly with the recurrence of liver metastasis. *GNG4* expression in a gastric cancer cell line was associated with malignant phenotypes, especially the formation of liver metastases.

The findings of our epidemiological data proved our expectation for *GNG4* as a biomarker in gastric cancer, especially in the context of liver metastasis. Higher expression of *GNG4* in primary gastric cancer tissue was significantly associated with poor prognosis caused mainly by liver recurrence. Additionally, the high *GNG4* group had less benefit from adjuvant chemotherapy aimed at preventing liver recurrence. The drug sensitivity test proved that *GNG4* is not only a candidate biomarker predicting resistance to chemotherapy, but may also have functional aspects of resistance to 5FU, a representative of pyrimidine drugs.

This study shows that *GNG4* is upregulated in malignant phenotypes, promoting cell survival and cell adhesion. This oncogenic phenotype of *GNG4* was much more apparent in a liver-metastatic mice model than a subcutaneous model. In the literature, *GNG4* is a member of the G protein γ family, which forms heterotrimers with the α and β subunits of G proteins. G proteins typically act as switches that transduce signals from upstream-acting G protein-coupled receptors (Crespo et al., 1994; Garcia-Regalado et al., 2008; Khan et al., 2013; Zhang et al., 2010). It is also reported that GNB1 and GNG2, family members of *GNG4*, independently maintain mitochondrial membrane polarity to evade apoptosis. We also detected *GNG4-*associated phosphorylation of ERK1/2, Akt, which are well-known oncogenic pathways promoting cell cycle, cell proliferation, and cell adhesion (Cheng et al., 2005; Garcia-Regalado et al., 2008; Khan et al., 2013). However, we are still unaware of published studies implicating *GNG4* as an oncogene.

Identification of *GNG4* as a candidate gene that accounts for liver metastases in gastric cancer can lead to a novel therapeutic or diagnostic strategy. If we recognize the risk of liver metastasis by testing on gastric cancer tissues, we may develop better disease monitoring weighted to the liver by MRI (Borggreve et al., 2019). Of note, most of the liver recurrence in this study occurred within six months to one year after surgery, implying the existence of subclinical dissemination to the liver at the time of surgery. Identifying such a cohort with higher risk may enable us to uncover further benefits from perioperative therapy. In particular, we may also seek a better chemotherapeutic approach targeted on liver metastasis through a clinical trial since some regimen reportedly may control hematogenous metastasis, rather than peritoneal metastasis, which is considered to have been controlled to some extent by regimens comprising fluorinated pyrimidines (Koizumi et al., 2008; Noh et al., 2014; Sakuramoto et al., 2007; Sasako et al., 2011). Moreover, discovering genes specific to liver metastasis, such as *GNG4*, may also help us to develop molecular-targeted drugs.

The present study has certain limitations. First, our *GNG4* mRNA expression data were retrospectively acquired, and extensive cohort prospective studies are required to validate this gene as a biomarker. Second, we may have to investigate the mechanisms of *GNG4*-mediated liver metastasis further. It may be proven through an organoid model of the sinusoid epithelium, for instance. Third, we have not shown how much specific *GNG4* drives liver metastases *in vivo* yet. Further studies using an orthotopic model may be needed to prove if *GNG4* expression affect organ directionality for cancer cells to metastasize (Busuttil et al., 2018).

Our results showed that higher *GNG4* expression correlated poorer prognosis, especially in the context of liver metastasis. We also proved that *GNG4* likely plays roles in promoting the cell cycle, evading apoptosis, and promoting tumor cell adhesion. *GNG4* may, therefore, represent a specific biomarker for detecting and putative therapeutic target for liver metastasis in patients with gastric cancer.

## Data Availability

Data is available in the manuscript and supplemental data. All other data will be available as needed unless related to personal information.

